# Evaluation of a Targeted Oral Microbiome Treatment in an Older Adult Population

**DOI:** 10.64898/2026.10.27.26366652

**Authors:** Hannah Moglowsky, Courtney Reynolds, Jenna Lovasz, Faheem M. Mir, Mausam Shah, Aavni Varadhan, Gabriella von Dohlen, Natalie Wall, Suparna A. Mahalaha, Lisa Blackmon, Sofronia M. Ringold, Farida Kassim Ejaz, Emily A. Stein

## Abstract

Gingivitis is highly prevalent among older adults and is associated with systemic diseases. Accessible and effective interventions are needed, particularly for underserved populations facing barriers to conventional dental care, such as older adults. This study evaluated the efficacy of a novel, multi-component lozenge (PTx800) designed to modulate the oral environment through blockade of sugar-to-acid and sugar-to-biofilm production of the oral microbiota. A six-week, double-blind, randomized, active placebo-controlled pilot trial was conducted with older adults (final sample n=20) residing in Cleveland, Ohio. Participants were randomized to receive either PTx800 or an active placebo lozenge containing sugar alcohols, used three times daily. Primary outcomes included Modified Gingival Index (MGI), Plaque Index (PI), Bleeding on Probing (BOP), Periodontal Probing Depth (PPD), and changes in oral pathogen load assessed by quantitative PCR. The PTx800 group demonstrated a 100% reduction in BOP, achieving complete resolution of bleeding sites, compared to an 82% reduction in the active placebo group. PPD was stabilized in the PTx800 group and slightly worsened in the control group. Microbiological analysis revealed a significant reduction in the total bacterial load in the PTx800 group, with pronounced decreases in key red complex periodontal pathogens, including *Porphyromonas gingivalis* (Pg), *Tannerella forsythia* (Tf), and *Treponema denticola* (Td). PTx800 lozenges significantly improved clinical and microbiological measures of gingival health in older adults compared to active placebo. These findings suggest that PTx800 is a promising, non-antibiotic approach for managing oral inflammation and dysbiosis in aging populations, with potential implications for reducing oral pathogens strongly linked to systemic diseases.

## INTRODUCTION

Gingivitis is an inflammatory condition characterized by swollen, red, and bleeding gums. It is the first stage of periodontal disease, the most common chronic inflammatory condition affecting aging populations. Severe periodontitis is the leading cause of tooth loss and impacts a significant percentage of adults over 65 years of age [1]. The burden of these diseases is not distributed equally. Older adults, particularly those from minority communities and lower socioeconomic backgrounds, face substantial barriers to accessing professional dental care, such as financial cost, lack of dental insurance, transportation difficulties, and a perceived lack of need [2]. These challenges, together with a high prevalence of tooth loss, infrequent dental visits, and numerous barriers to care, underscore the urgent need for accessible, affordable, and effective interventions that can be implemented outside of a traditional care setting.

The consequences of untreated oral inflammation extend far beyond the oral cavity. A robust body of evidence has established the “oral-systemic link,” wherein dysbiosis of the oral microbiome is implicated in the pathogenesis and progression of numerous systemic diseases prevalent in aging [3]. Periodontal pathogens and their inflammatory byproducts can enter the bloodstream, and contribute to conditions such as cardiovascular disease, type 2 diabetes, respiratory disease, and neurodegenerative disorders including Alzheimer’s disease [4]. A central actor in this process is *Porphyromonas gingivalis* (Pg), a keystone pathogen that not only drives local periodontal ligament destruction but also acts as a “conductor” to orchestrate systemic inflammation [5]. The ability of Pg and its virulence factors to translocate to distal sites highlights the oral cavity and subgingival mucosa as a critical reservoir of pro-inflammatory anaerobes [6].

Current management strategies for gingivitis and periodontitis rely primarily on mechanical biofilm disruption (scaling and root, debridement) and the use of antiseptics and antibiotics. While these may provide a temporary reduction in pathogen burden, they are not curative. Systematic reviews show an average 9% reduction in BOP using conventional approaches to biofilm disruption [7]. In addition, antimicrobials can pose risks with long-term or frequent use [8]. The increasing threat of antimicrobial resistance (AMR) necessitates the development of novel non-antimicrobial approaches [9]. PTx800 was identified from high-throughput screening of Generally Recognized As Safe (GRAS) ingredients on American Type Culture Collection (ATCC) dental caries and periodontitis biofilm isolates. Compounds were categorized into sugar-to-acid blockade, sugar-to-biofilm blockade, selective disruption of pathogen growth, and preservation of commensal growth. PTx800, a dual ingredient mixture of cyclodextrin and vitamin B6, was selected for its superior activity in sugar blockade and selective growth activities with invitro biofilm formation and sugar challenge growth assays.

This study aimed to evaluate the efficacy of PTx800 in reducing clinical and microbiological markers of gingivitis in a population of older adults drawn from a county that included senior centers serving clients in urban, low-income neighborhoods as well as more affluent suburban centers. The primary hypotheses were that, compared to an active placebo containing sugar alcohols, the PTx800 lozenge would lead to a statistically significant reduction in clinical signs of gingival inflammation and bacterial counts of keystone periodontal pathogens.

## MATERIALS AND METHODS

### Study Design and Ethical Approval

A six-week, double-blind, randomized, active placebo-controlled experimental study was conducted in partnership with the Benjamin Rose Institute and its senior centers, i.e., the Rose Centers for Aging Well (RCAW). This study was registered on ClincalTrials.gov (NCT03693027; Registration date October 2, 2018). The study protocol was approved by the Institutional Review Board of the Benjamin Rose Institute on Aging and adhered to the ethical principles for medical research involving human subjects outlined in the Declaration of Helsinki [10]. All participants provided written informed consent prior to enrollment, and all data were anonymized to maintain confidentiality.

### Participant Recruitment and Screening

Participants were recruited from July to October 2018. Enrollment was completed in December 2018. Two phases of the study were designed. In the first phase, a survey was conducted with 122 senior center participants regarding their oral health habits/hygiene, perceptions of dental health and dental visits including barriers to dental care. The focus of this paper is on the second phase of this study, i.e., the randomized clinical trial to test the efficacy of PTx800 in reducing clinical and microbiological markers of gingivitis in an underserved population of older adults. The eligibility criteria for the second phase were divided into three steps. In the first step, investigators excluded participants who had legal guardians, used tobacco within the past two years, and had full dentures. Of the 122 participants from Phase I, 52 met the initial eligibility criteria for the study; however, only one participated.

Additional participants were recruited from senior centers across northeast Ohio via flyers and outreach events. Interested respondents completed the Short Portable Mental Status Questionnaire (SPMSQ), a validated cognitive screening tool assessing orientation, memory, and attention; individuals with five or more errors were excluded [11]. Those who passed the cognitive screen underwent an on-site dental screening.

In total, 230 individuals were contacted, of which 141 met the criteria for study participation, prior to the dental screening. Of these, 67 were deemed ineligible after the dental examination, leaving a potential sample of 74 older adults. Of the 74, 47 older adults either passively or verbally refused to participate, and 27 completed the study for a response rate of 36%. The on-site screening began with an initial chairside visual examination by a dental professional to determine general oral health status. Participants who passed this initial check proceeded to a comprehensive dental assessment. Inclusion criteria for the interventional stage included: a) aged 60 years or older, b) no legal guardian, c) non-smoker for at least two years, d) mild-to-moderate gingivitis (defined as a Modified Gingival Index score of ≥1.75), e) at least 12 natural teeth, f) no partial or full dentures, and g) self-reported bleeding gums upon brushing. Exclusion criteria included: a) severe periodontitis, b) gross caries, c) significant oral soft tissue pathology, d) use of antibiotics within 30 days prior to enrollment, or e) participation in another dental clinical trial within the previous 30 days.

### Randomization and Intervention

Participant randomization was generated using a computer software program in a 1:1 allocation sequence. The randomization sequence was generated by a person not involved in recruitment or clinical evaluations. The coordinating site received one package containing 60 individual envelopes identified by participant number. Sequence concealment was maintained using sequentially numbered, opaque, sealed envelopes. Each sealed envelope contained the assigned intervention details (lozenge name, formulation, and manufacturer lot number). To maintain blinding for both participants and the study coordinator, the bottle labeling for both active and placebo lozenges was coded. Neither the personnel enrolling participants, the personnel assigning interventions, the dental examiner, nor the study participants had access to the master randomization sequence or knowledge of intervention assignments throughout the trial.

The sample of 27 eligible participants were randomized to the PTx800 group (intervention group, n=15) or the active placebo lozenges (control group, n=12). The PTx800 lozenge (manufactured by Solara Labs, Miami Gardens, FL) contained a proprietary blend of cyclodextrin, pyridoxal-5-phosphate, calcium carbonate, and sodium bicarbonate, along with inactive ingredients (mannitol, sorbitol, modified cellulose, natural mint flavor, magnesium stearate, stevia leaf extract). The active placebo lozenge (manufactured by Solara Labs, Miami Gardens, FL) contained sugar alcohol and other inactive ingredients but lacked the primary active components. Both lozenges were manufactured under cGMP as mandated by the US FDA and were identical in appearance and taste. Participants, study personnel, and the testing laboratory were blinded to the treatment allocation.

### Study Procedure and Participant Adherence to Intervention

Study visits occurred at baseline (Visit 1), three weeks (Visit 2), and six weeks (Visit 3). At Visit 1 (baseline) and Visit 3 (post-test), participants were surveyed about their oral health conditions and habits, physical and mental health, and well-being. This study focuses on the clinical oral health assessments, including MGI, PI, BOP, and PPD measurements that were performed during each visit by a licensed dental professional. Oral rinse samples were also collected at each visit.

Participants were instructed to let one lozenge dissolve in their mouth for 60-120 seconds after each meal, three times per day, for six weeks. They were asked to maintain their usual oral hygiene habits but abstain from using mouthwash. Adherence was monitored via self-reported daily logs, and lozenge counts at each visit; consumption of ≥70% of assigned lozenges was required for inclusion in the final analysis. Participants were contacted periodically over the duration of the study to address any questions or concerns. Adverse events were systematically assessed at each visit.

### Clinical Oral Health Assessments Modified Gingival Index

Modified Gingival Index (MGI) scoring is a visual assessment used to quantify the level of gingival inflammation [12]. The scoring system rates mild and moderate inflammation where 0=absence of inflammation, 1=mild inflammation or with slight changes in color and texture but not in all portions of gingival marginal or papilla, 2=mild inflammation, such as the preceding criteria, in all portions of gingival marginal or papilla, 3=moderate, bright surface inflammation, erythema, edema and/or hypertrophy of gingival marginal or papillary, and 4=severe inflammation of erythema, edema and/or marginal gingival hypertrophy of the unit or spontaneous bleeding, congestion or ulceration. The MGI is obtained by adding the values of each tooth and dividing by the number of teeth examined. A score from 0.1-1.0 indicates mild inflammation, 1.1-2.0 indicates moderate inflammation, and 2.1-3.0 indicates severe inflammation.

### Plaque Index

The Turesky Plaque Index (PI) measures plaque accumulation using a disclosing solution and is scored on a 0 to 5 scale in which a score of 0=no plaque, 1=separate flecks of plaque, 2=continuous band of plaque less or equal to 1mm, 3=continuous band of plaque greater than 1mm but less than 1/3 of crown height, 4=continuous band of plaque greater than or equal to 1/3 but less than or equal to 2/3 of crown height, and 5=continuous band of plaque greater than 2/3 of crown height. Each tooth received six individual scores at mesial, middle, and distal aspect for both the facial and lingual surfaces. Each participant’s score was calculated by adding the scores at each site and dividing by the number of sites evaluated. Higher scores denote greater plaque accumulation [13].

### Bleeding on Probing

Bleeding on Probing (BOP) was assessed using a periodontal probe with a 0.5 mm diameter tip inserted into the gingival crevice, swept from distal to mesial around the tooth at a depth of approximately 1mm and at an angle of approximately 60° while in contact with the sulcular epithelium. Each of 4 gingival areas (disto-buccal, midbuccal, mesio-buccal and mid-lingual) around each tooth was assessed in this manner waiting approximately 30 seconds before recording the number of gingival units which bleed, according to the following scale: 0=absence of bleeding after 30 seconds, 1=bleeding observed after 30 seconds, and 2=immediate bleeding observed. The number of bleeding sites was used to calculate the gingival bleeding score [14].

### Periodontal Probing Depth

Periodontal Probing Depth (PPD), also known as pocket depth, was measured as the distance in millimeters from the gingival margin to the base of the pocket at six sites per tooth/implant using standardized disposable double-ended explorer periodontal probes (Practicon, Greenville, NC) calibrated to 1mm increments [15,16]. Measurements were obtained by inserting the probe parallel to the root surface along the periodontal pocket axis until slight resistance was met at the base of the pocket, following established periodontal assessment protocols. The deepest measured value per anatomical section (mesial, mid, distal) was recorded for clinical evaluation.

### Sample Collection

Oral rinses were collected from participants at baseline and six weeks. Participants were instructed to rinse thoroughly with 4mL of sterile water for 15 seconds, after which the rinse was expelled into a sterile collection tube. All samples were immediately labeled and time stamped. Samples were stored at 4°C at the study site before being transported on ice to Oral DNA Labs for testing within 24 hours. Processing and analysis were performed by Oral DNA Labs according to FDA registered test methods for MyPerioPath® qPCR test (Oral DNA Labs, Eden Prairie MN).

### Bacterial Oral Pathogens

Periodontal pathogen quantification via the MyPerioPath® qPCR test (Oral DNA Labs, Eden Prairie MN) was performed in a CAP-accredited and CLIA-certified laboratory. This laboratory-developed test followed internally validated protocols compliant with CLIA analytical validity requirements (42 CFR §493.1253) and CAP checklist standards (MIC.22750). The following periodontal pathogens were assessed at baseline and week six from oral rinse samples: *Aggregatibacter actinomycetemcomitans* (Aa), *Campylobacter rectus* (Cr), *Capnocytophaga* spp. (*gingivalis, ochracea, sputigena*; (Cs), *Eikenella corrodens* (Ec), *Eubacterium nodatum* (En), *Fusobacterium nucleatum* (Fn), *Peptostreptococcus (Micromonas) micros* (Pm), *Porphyromonas gingivalis* (Pg), *Prevotella intermedia* (Pi), *Treponema denticola* (Td), and *Tannerella forsythia* (Tf). Bacteria levels are reported in bacterial colony-forming units per mL (CFU/mL) of oral rinse sample.

### Assessment of Outcomes

Primary outcomes for the clinical trial included changes in clinical oral health measures (MGI, PI, BOP, PPD) and reductions in oral pathogen burden. Secondary outcomes included general health and lifestyle factors.

### Clinical Outcomes Analysis

All statistical analyses were performed using R statistical software *version 4.4.2* (R Foundation for Statistical Computing, Vienna, Austria). Baseline characteristics were compared between the PTx800 and control groups using Mann-Whitney U tests for continuous variables (MGI, PI, BOP, PPD and microbiological CFU counts) and chi-square tests for categorical variables (e.g., gender, smoking status and treatment group). To assess relationships within the dataset, Kendall’s rank correlation coefficients were used to evaluate associations between clinical measures and pathogen concentrations (CFU counts). The Difference-in-Differences (DiD) method was applied to compare temporal changes between cohorts for each pathogen using a linear model at baseline and six-week timepoints. For microbiological data, CFU counts for pathogens, including Aa, Cr, Cs, Ec, En, Fn, Pm, Pg, Pi, Td and Tf were log-transformed before analysis to normalize distributions. Participants with CFU levels of zero at both baseline and endpoint were excluded from pathogen-specific analyses. Mann-Whitney tests were used to compare CFU counts and relative abundances within and between cohorts. For pocket depth, statistical comparisons were conducted both using per-person averages across all measured teeth and using all individual tooth measurements. Effect sizes were calculated using Cohen’s d formula for statistically significant findings (p < 0.05). Missing data were handled using multiple imputation, consistent with the study protocol. All statistical tests were two-sided, and an alpha level of 0.05 was used to assess significance.

## RESULTS

### Participant Flow and Baseline Characteristics

The final cohort had a mean age of 71 years (range 57-86). Most participants were female (72%) and identified as either Black or African American (44%) or White (41%). Over the six-week intervention, one participant withdrew prior to randomization, three participants withdrew from the control group and four from the PTx800 group. The final analysis included 20 participants, 11 in the PTx800 group and 9 in the control group (Figure 1). The two groups were well-matched at baseline with respect to demographic characteristics and clinical oral health measures, with no statistically significant differences observed (Figure 2A-B). There was one adverse event that was categorized as mild severity and unlikely related to participation in the study. During the study, one participant reported experiencing transient lightheadedness and headache after consuming a lozenge and subsequently chose to withdraw. The participant noted a prior history of similar episodes and concurrent use of multiple antihypertensive medications. The Principal Investigator determined the event to be mild in severity (Grade 1) and unlikely to be related to the study product. The event resolved spontaneously without requiring medical treatment, and no changes to study protocols or consent documentation were necessary.

**Figure 1.**
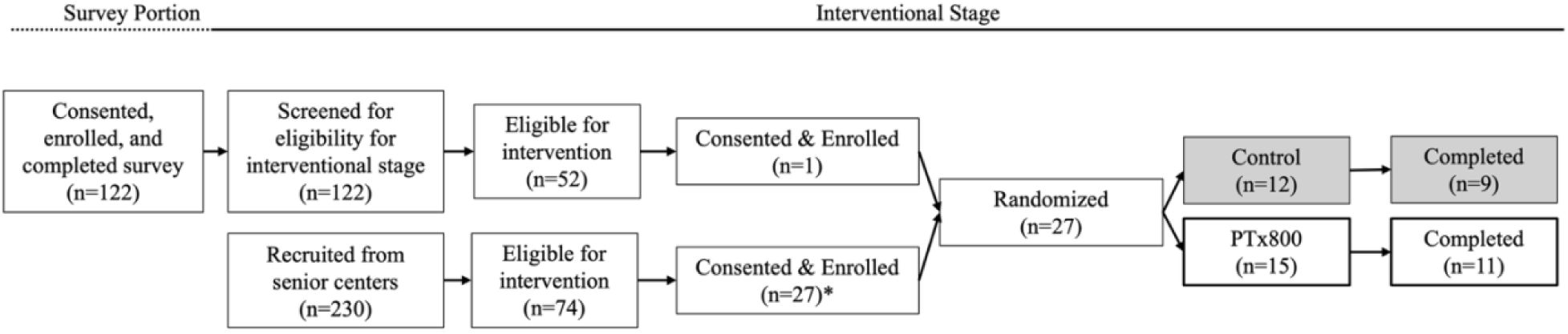
Study Flow Diagram. Flowchart illustrating participant enrollment, randomization, and study completion. Participants were initially enrolled in a survey or recruited more broadly for specifically the interventional portion of the study after which eligible individuals proceeded to the interventional stage. Randomized participants were assigned to either the Control or PTx800 group. *: One participant withdrew prior to randomization.

**Figure 2.**
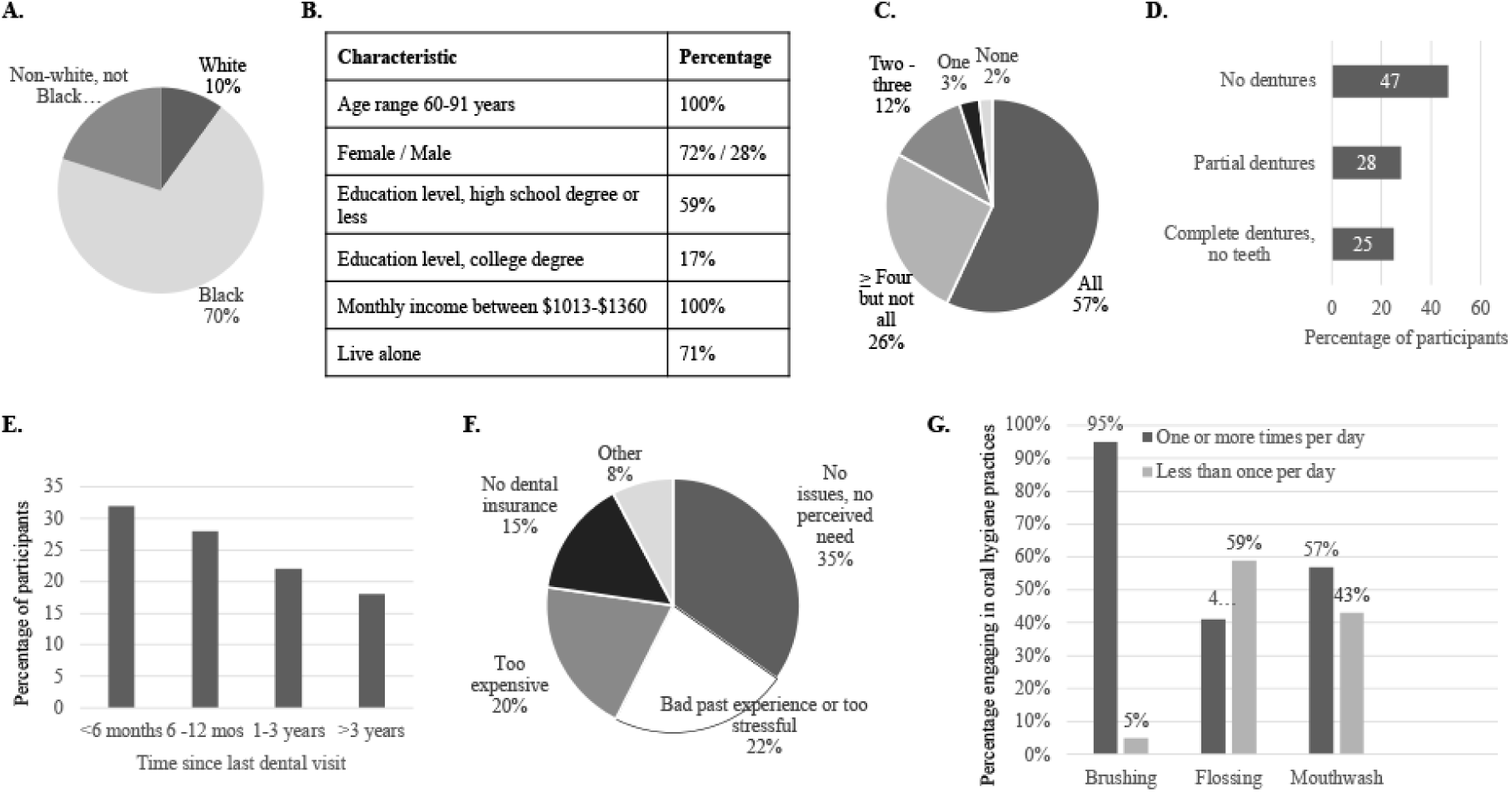
Demographic characteristics and oral health-related factors of study participants. (**A**) Racial distribution of study participants. (**B**) Key demographic characteristics, including age range, gender distribution, education level, income, and living status. (**C**) Proportion of participants with tooth loss. (**D**) Use of dentures among participants. (**E**) Time since last dental visit. (**F**) Reported barriers to accessing dental care. **(G)** Frequency of daily oral hygiene behaviors among participants, including brushing, flossing, and mouthwash use. Data are expressed as percentages of total participants.

### Socio-behavioral Profile and Barriers to Oral Healthcare

Participants reported a high frequency of daily tooth brushing (95%), but less consistent flossing (41% daily) and mouthwash use (57% daily) (Figure 2G). Access to professional dental care was limited; 33% had visited a dentist in the last six months, and 18% had not had a dental visit in over three years (Figure 2E). The primary self-reported barriers to care were a perceived lack of need (35%), financial cost (20%), and a lack of dental insurance (15%) (Figure 2F).

### Disparities in Baseline Systemic Comorbidities

Despite successful randomization on demographic and primary clinical variables, significant imbalances in self-reported systemic health conditions were present between the groups at baseline (Figure 3A). The PTx800 group reported a substantially higher prevalence of hypertension (93% vs. 40%), arthritis (79% vs. 50%), acid reflux/GERD (43% vs. 9%), and dry mouth (50% vs. 30%) compared to the control group (Figure 3B). Conversely, the control group reported a higher prevalence of kidney disease (32% vs. 7%) and thyroid problems (23% vs. 7%), data not shown. These pre-existing differences were noted but not adjusted for in the primary outcome analysis.

**Figure 3.**
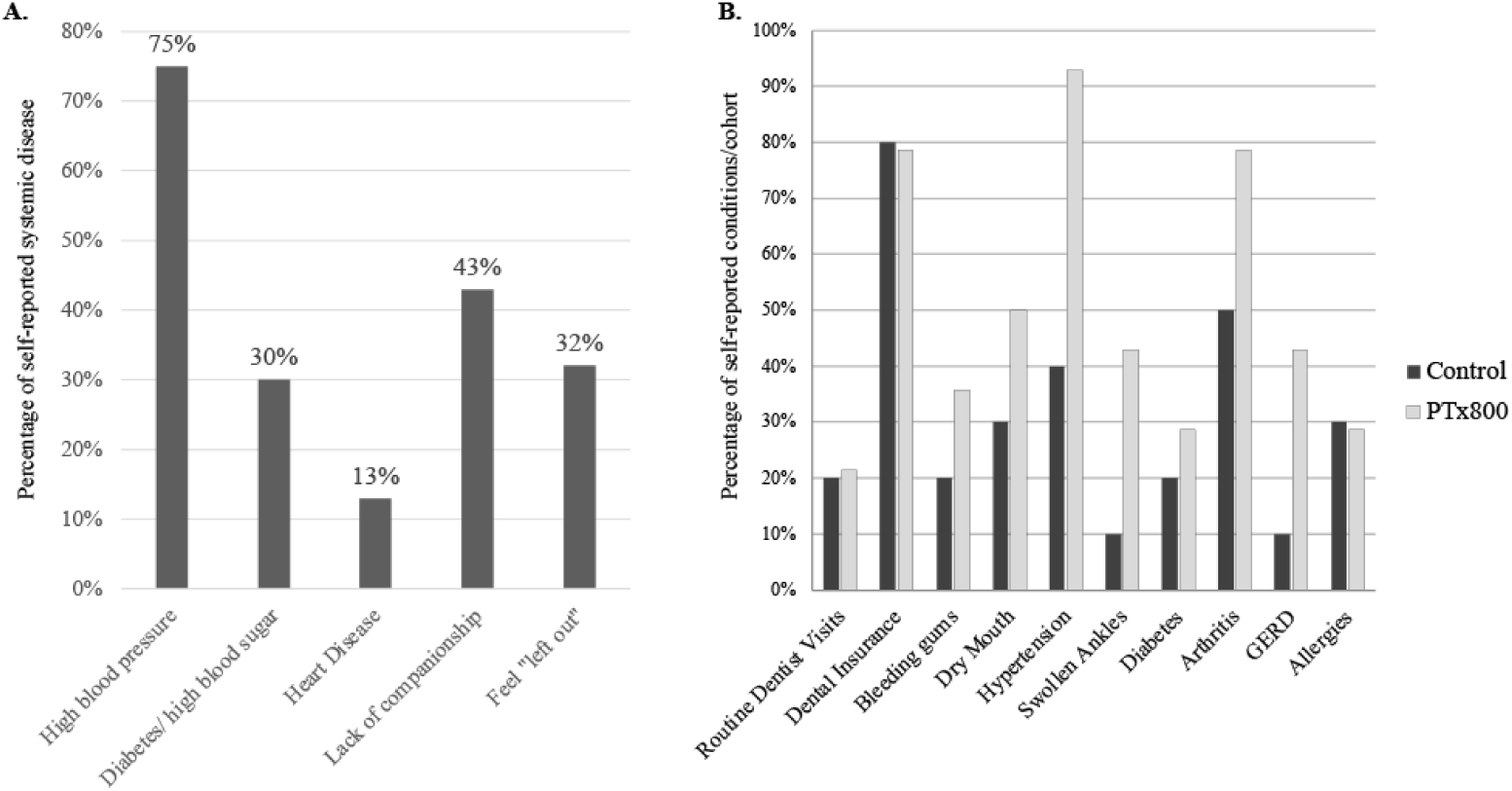
Baseline Medical and Dental Characteristics. **(A)** Self-reported health conditions and social factors potentially influencing oral health behaviors from the 127 participants in the survey portion of the study. Data are expressed as percentages of total participants. **(B)** Prevalence of self-reported medical and dental conditions among participants in the interventional stage of the study, stratified by treatment group (Control vs. PTx800). Data are expressed as percentages of participants in each group.

### PTx800 Lozenge Intervention Significantly Ameliorates Clinical Signs of Gingival Inflammation

After six weeks of intervention, participants in the PTx800 group showed superior improvements in key clinical measures of gingival health compared to the control group. The most pronounced effect was observed in BOP. The PTx800 group experienced a 100% mean reduction in bleeding sites, indicating a complete resolution of active gingival inflammation. In contrast, the control group also showed a substantial but incomplete improvement, with an 82% reduction in BOP (Figure 4C). The difference in improvement between the groups was statistically significant (p = 0.04). For PPD, the groups showed divergent trends. The PTx800 group demonstrated a slight average decrease in pocket depth (−0.47%), suggesting stabilization or modest improvement of periodontal health. The control group, however, experienced a slight average increase (+0.19%), potentially indicating a subtle progression of disease severity over the study period (Figure 4G). Both groups showed similar modest reductions in MGI and PI scores. The PTx800 group had a 36% reduction in MGI and a 29% reduction in PI, while the control group had a 39% reduction in MGI and a 27% reduction in PI scores (Figure 4A, Figure 4E). These improvements were not statistically different between the groups.

**Figure 4.**
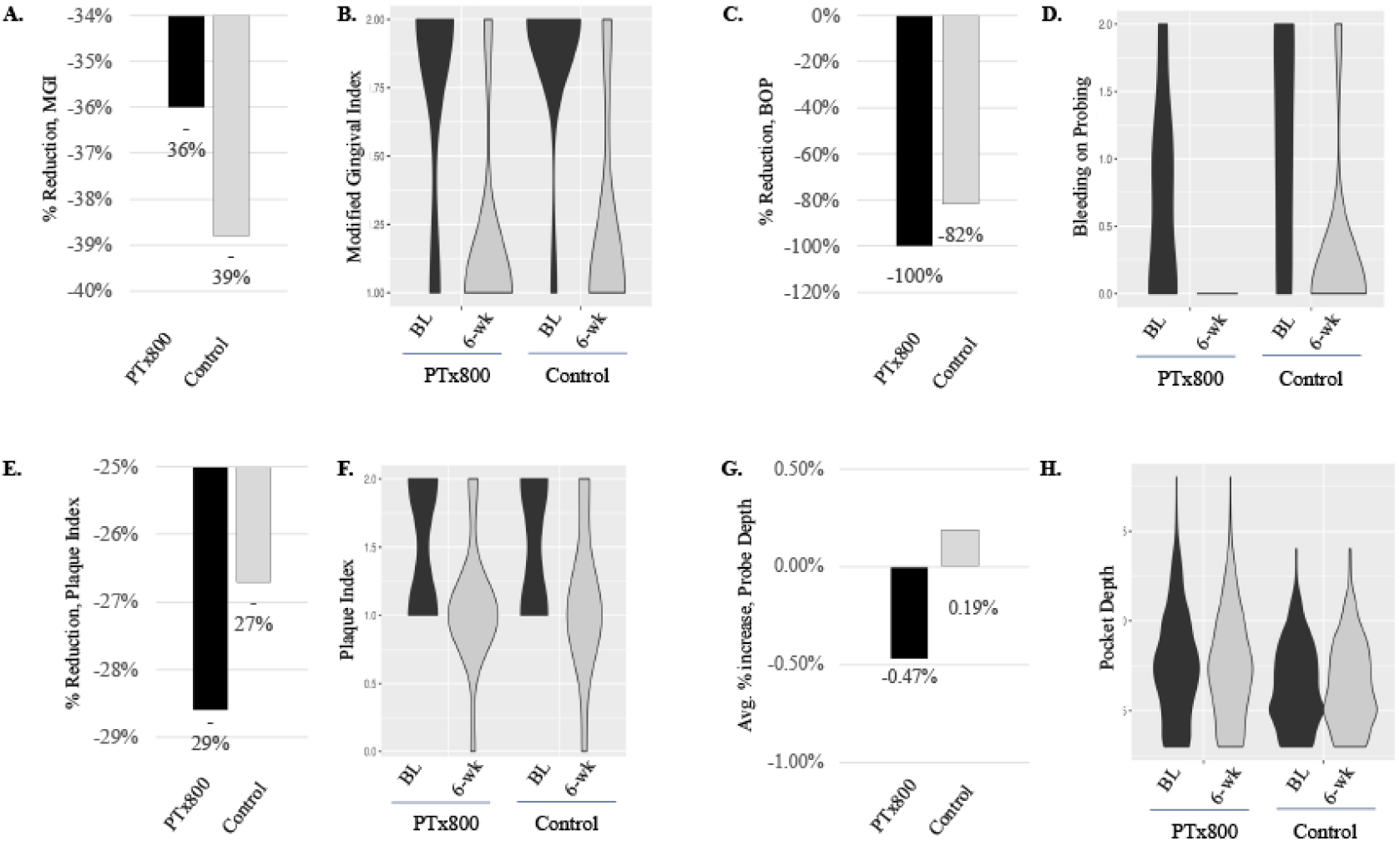
Changes in clinical oral health measures over six weeks of lozenge use. Bar plots illustrate average percent changes from baseline to six-week endpoints in clinical measures among participants in the PTx800 and control groups: average percent reduction in (A) MGI, (C) BOP, and (E) PI, and average percent change in (G) periodontal probing depth. Negative values represent reductions, positive values represent increases. Violin plots illustrate data distribution and density of clinical scores at baseline (black) and six weeks (gray) in both cohorts for (**B**) MGI scores, (**D**) number of bleeding sites assessed by BOP, (**F**) PI scores, and (**H**) periodontal probing depth measurements. Wider sections indicate higher participant density at a given score, while narrower plot areas indicate fewer participants at corresponding clinical scores.

### PTx800 Modulates the Oral Pathogen Burden with a Pronounced Reduction in Red Complex Bacteria

Quantitative PCR analysis revealed that the PTx800 intervention led to a marked reduction in the oral burden of key periodontal pathogens, whereas the control group exhibited minimal change. The overall bacterial load (total CFU/mL) decreased in the PTx800 group. Most notably, the PTx800 group showed significant reductions in the red complex pathogens, which are strongly associated with destructive periodontitis. Levels of *P. gingivalis*, *T. forsythia*, and *T. denticola* all decreased substantially from baseline to week six in the intervention group. Additionally, the pathobiont *E. corrodens* was also reduced. In contrast, levels of these pathogens remained stable or increased slightly in the control group. *A. actinomycetemcomitans*, a pathogen associated with aggressive periodontitis, was detected at baseline only in the PTx800 group and was undetectable in all PTx800 participants by the six-week endpoint (Figure 5).

**Figure 5.**
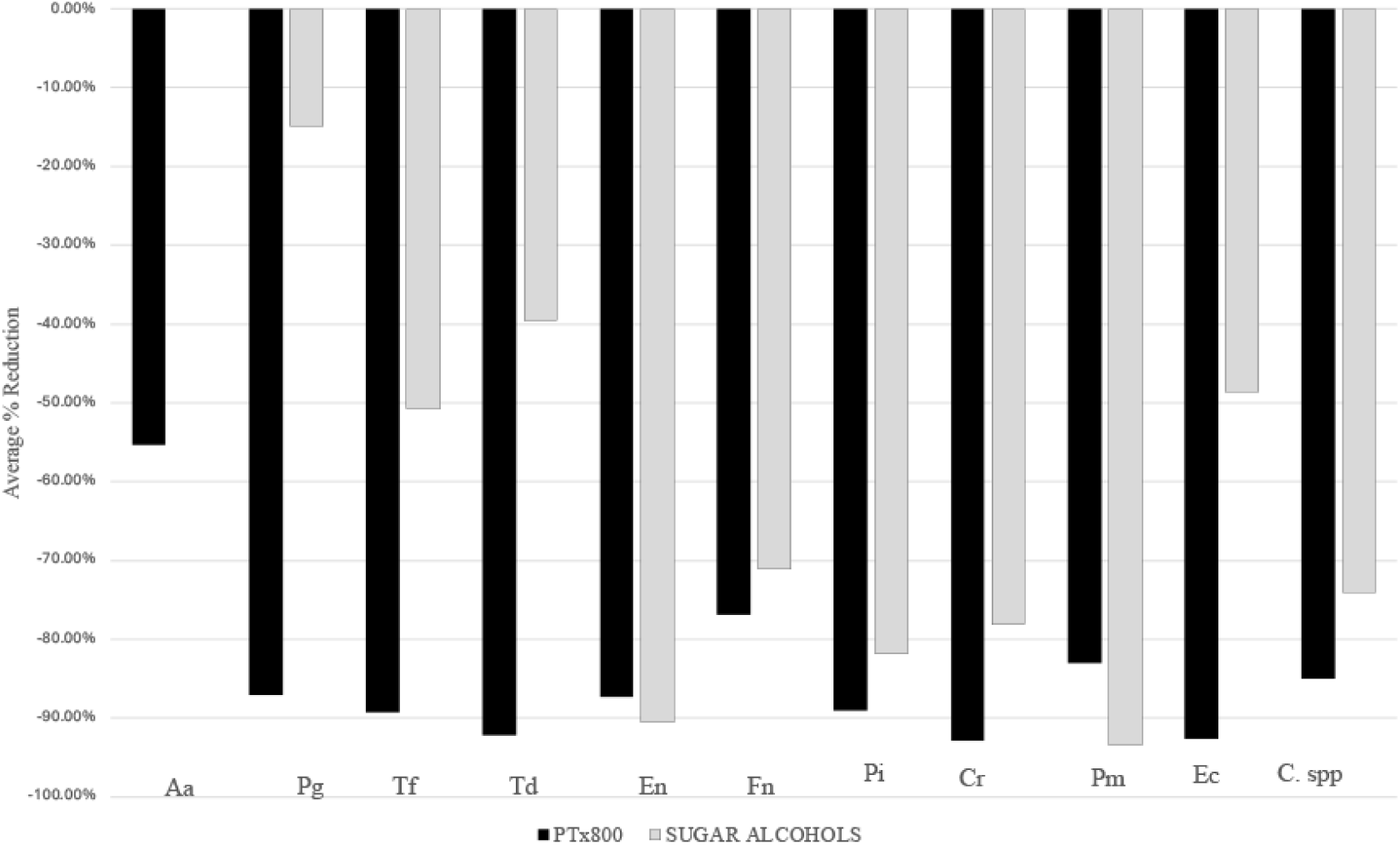
Reduction in bacterial load after six weeks of lozenge use. Percent reduction in colony-forming units per milliliter (CFU/mL) for oral pathogens in the PTx800 (black) and control (grey) cohorts, measured using the MyPerioPath® quantitative PCR test. Bacterial species analyzed include *Aggregatibacter actinomycetemcomitans* (Aa), *Porphyromonas gingivalis* (Pg), *Tannerella forsythia* (Tf), *Treponema denticola* (Td), *Eubacterium nodatum* (En), *Fusobacterium nucleatum* (Fn), *Prevotella intermedia* (Pi), *Campylobacter rectus* (Cr), *Peptostreptococcus micros* (Pm), *Eikenella corrodens* (Ec), and *Capnocytophaga* spp. (C. spp.). *A. actinomycetemcomitans* (Aa) was not detected in any control samples at baseline or endpoint. Negative values indicate a reduction in bacterial load relative to baseline.

## DISCUSSION

This randomized controlled pilot trial provides preliminary evidence that PTx800 lozenges improve clinical and microbiological markers of gingival health in older adults over a six-week intervention period. The most striking finding was the complete elimination of BOP in the PTx800 group – a 100% reduction compared to an 82% reduction in the active placebo control group. This difference was statistically significant (p=0.04) and clinically meaningful, as BOP serves as a sensitive indicator of active gingival inflammation and is strongly predictive of periodontal disease progression [14,17,18]. The achievement of complete BOP resolution in six weeks is noteworthy given that this cohort consisted of community-dwelling older adults with pre-existing gingivitis, many of whom reported systemic health conditions that typically complicate oral health management.

### Clinical Significance of Bleeding on Probing

The superior performance of PTx800 in eliminating BOP represents a substantial clinical advancement for managing gingivitis in older adults. BOP reflects acute inflammation within the gingival sulcus and is considered one of the most reliable early indicators of periodontal tissue destruction [17,18]. The complete resolution observed in the PTx800 group suggests that the intervention not only reduced surface-level inflammation but also addressed underlying inflammatory processes at the tissue level. This is particularly significant because chronic gingival inflammation, if left untreated, can progress to periodontitis—a condition affecting a substantial proportion of older adults that has been linked to numerous systemic health complications [19,20].

In contrast to BOP, which reflects acute inflammation, the Modified Gingival Index captures more chronic, visual signs of inflammation and showed only modest reductions in both groups (PTx800 36%, Control 39%). This discrepancy between BOP and MGI improvements is expected and informative. BOP is a more sensitive measure of deep tissue inflammation and vascular permeability changes that often precede visible alterations in gingival appearance. The more pronounced improvement in BOP compared to MGI suggests that PTx800’s therapeutic effects manifest first at the vascular and inflammatory level before producing changes in gross gingival morphology [14,21]. A longer study duration may be required to observe significant improvements in MGI scores as chronic inflammation resolves and the tissues undergo remodeling.

The probing depth measurements provide additional context for interpreting these clinical improvements. Periodontal probing is a widely studied method for assessing disease status, with research having established the relationship between probe penetration and underlying clinical and histologic signs of inflammation [16,21,22]. The PTx800 group demonstrated a slight average decrease in pocket depth (−0.47%), suggesting stabilization or modest improvement of periodontal health, while the control group experienced a slight average increase (+0.19%), potentially indicating a subtle progression of disease severity over the study period. Although both changes were modest, the divergent trends are clinically relevant—particularly in an aging population where progressive pocket deepening often signals advancing periodontitis [23]. The stabilization achieved by PTx800 is especially meaningful given that many participants in this group had baseline systemic conditions, such as arthritis and xerostomia, that typically impair oral hygiene practices and accelerate periodontal disease progression [19,20].

### Microbiological Impact: Targeting the Red Complex Pathogens

The clinical improvements in the PTx800 group were accompanied by profound shifts in the oral microbiome composition. Quantitative PCR analysis revealed that PTx800 markedly reduced the load of the red complex pathogens (Pg, Tf, Td) which are strongly associated with destructive periodontitis and considered keystone pathogens in periodontal disease progression [8,24,25]. These anaerobic bacteria colonize subgingival plaque biofilms, produce potent virulence factors including proteolytic enzymes and lipopolysaccharides, and orchestrate dysbiotic microbial communities that drive chronic inflammation and tissue destruction [5,26]. The reduction of these pathogens from baseline to week six in the PTx800 group, while levels remained stable or increased slightly in the control group, demonstrates PTx800’s capacity to modulate the pathogenic oral microbiome toward a healthier, more balanced state.

Of particular significance is the reduction in *Porphyromonas gingivalis*, which serves as a biomarker of gingival and periodontal infection and represents one of the most extensively studied periodontal pathogens. *P. gingivalis* possesses unique virulence characteristics that enable it to evade host immune responses, manipulate complement pathways, and persist in subgingival biofilms despite the host’s inflammatory response [5,24]. Beyond the oral cavity, accumulating evidence implicates *P. gingivalis* in the pathogenesis of several systemic disorders, most notably Alzheimer’s disease and cardiovascular disease [27]. Studies have demonstrated that *P. gingivalis* can translocate from periodontal tissues to distant anatomical sites including the cerebrospinal fluid, brain parenchyma, coronary arteries, and liver [28,29]. The bacterium’s gingipain proteases have been detected in the brains of Alzheimer’s disease patients, and experimental models demonstrate that oral infection with *P. gingivalis* can induce neuroinflammation, amyloid-beta deposition, and cognitive impairment [29]. Similarly, *P. gingivalis* DNA and viable bacteria have been identified in atherosclerotic plaques, and periodontal infection has been associated with increased cardiovascular disease risk [22,30].

The demonstrated ability of PTx800 to reduce *P. gingivalis* burden therefore carries implications that extend far beyond oral health. Given the growing recognition that periodontal pathogens—particularly *P. gingivalis*—may contribute to systemic disease pathogenesis, interventions that effectively reduce these bacteria in the oral cavity could potentially mitigate their systemic dissemination and associated disease risks. This possibility is especially relevant for older adults, who experience higher rates of both periodontal disease and systemic conditions such as Alzheimer’s disease and cardiovascular disease [29–31]. Further research is needed to determine whether sustained reduction of *P. gingivalis* can reduce biomarkers of systemic inflammation, slow cognitive decline in at-risk populations, or improve cardiovascular outcomes.

In addition to the red complex organisms, PTx800 also reduced *Eikenella corrodens*, a pathobiont found in subgingival plaque that has been associated with various oral and systemic infections [32,33]. The intervention also eliminated *Aggregatibacter actinomycetemcomitans* (Aa), a pathogen associated with aggressive periodontitis, from the PTx800 group where it was uniquely detected at baseline [34]. By the six-week endpoint, Aa was non-detectable in all PTx800 participants. Together, these findings suggest that PTx800 helps re-establish microbial balance by selectively reducing key pathogens and pathobionts that thrive in dysbiotic biofilm communities, particularly those residing in difficult-to-access subgingival niches. However, due to the small sample size, these preliminary findings should be interpreted with caution and warrant validation in larger, tong-term clinical trials.

### Proposed Mechanisms of Action

The mechanisms underlying PTx800’s clinical and microbiological effects likely involve multiple complementary pathways. PTx800 contains a proprietary blend of botanical extracts and bioactive compounds that have demonstrated antimicrobial, anti-inflammatory, and immunomodulatory properties in previous studies. The formulation’s ability to reduce red complex pathogens suggests direct or indirect antimicrobial activity against these anaerobic species, potentially through disruption of biofilm architecture, interference with bacterial adhesion, or modulation of the subgingival environment to favor commensal over pathogenic species. Unlike broad-spectrum antibiotics, which face growing challenges with resistance development—including among periodontal pathogens like *P. gingivalis*—PTx800 appears to work through alternative mechanisms involving nutrient stress, that may be less susceptible to resistance evolution [35]. This is particularly important given that *P. gingivalis* shows a growing propensity for antibiotic resistance, necessitating interventions that can modulate microorganisms through non-antibiotic means [8].

The lozenge delivery format may enhance PTx800’s efficacy by promoting sustained contact between active ingredients and oral tissues. As the lozenge dissolves slowly over several minutes, bioactive compounds are distributed throughout the oral cavity via saliva, potentially reaching subgingival sites that are difficult to access with conventional oral hygiene practices [25,36,37]. The formulation may also contribute to the observed reduction in BOP by dampening the host’s overactive immune response to bacterial challenge, thereby reducing vascular permeability and tissue damage while allowing immune-mediated clearance of pathogens to proceed [14,19,21].

### Active Placebo Design and Control Group Improvements

A unique and important aspect of this study’s design was the use of an active placebo control comprised entirely of sugar alcohols (xylitol, sorbitol, and erythritol). This represents a rigorous test of PTx800’s efficacy, as sugar alcohols themselves possess documented oral health benefits [38]. Xylitol in particular has been shown to inhibit *Streptococcus mutans* growth, reduce plaque formation, and enhance remineralization. Sugar alcohols are non-fermentable by most oral bacteria, thus creating an unfavorable environment for acidogenic species, and may exert mild antimicrobial effects [39]. The substantial improvements observed in the control group—including an 82% reduction in bleeding on probing and approximately 40% reduction in gingival inflammation—attest to the therapeutic activity of the sugar alcohol base.

Furthermore, using a lozenge three times daily stimulates saliva flow, which provides multiple oral health benefits including mechanical cleansing of bacterial debris, acid buffering, delivery of antimicrobial components (lysozyme, lactoferrin, immunoglobulins), and maintenance of the oral mucosal barrier [40–42]. Enhanced saliva flow is particularly beneficial for older adults, many of whom experience medication-induced xerostomia that predisposes them to increased plaque accumulation and gingival inflammation [43]. Additionally, the “Hawthorne effect,” whereby study participation itself motivates participants to improve their oral hygiene behaviors, likely contributed to improvements in both groups [44].

PTx800 significantly surpassed the combined benefits of sugar alcohols, saliva stimulation, and enhanced hygiene motivation, underscoring its specific therapeutic action. The study results demonstrate an unequivocal benefit from the PTx800 bioactive ingredients, validating the formulation’s therapeutic rationale and suggesting that the observed effects cannot be attributed solely to the delivery vehicle or study participation effects.

### Oral Health in the Aging Populations

This study addresses an important public health concern: the oral health needs of the aging population. Older adults face multiple challenges to maintain optimal oral health, including age-related declines in manual dexterity, reduced saliva flow, comorbid systemic conditions, limited access to dental care, and cumulative lifetime exposure to oral diseases [43]. Gingivitis and periodontitis affect a substantial proportion of older adults, with prevalence increasing with age. The National Health and Nutrition Examination Survey has consistently documented that severe periodontitis affects approximately one in five adults aged 65 and older in the United States, with even higher rates among certain demographic subgroups [45].

The burden of periodontal disease in older adults is compounded by its bidirectional relationships with systemic health conditions that are highly prevalent in this age group. Periodontal disease has been associated with cardiovascular disease, diabetes, respiratory diseases, various cancers, rheumatoid arthritis, and cognitive decline including Alzheimer’s disease [22,29,46–48]. Interventions that effectively manage gingival inflammation and periodontal pathogen burden in older adults may yield benefits that extend beyond oral health, potentially influencing systemic inflammatory status and chronic disease trajectories.

PTx800 represents a particularly appropriate intervention for older adults because it is delivered as a lozenge that requires no specialized technique, minimal manual dexterity, and can be easily integrated into daily routines. This accessibility is crucial for older adults with arthritis or other conditions that impair the fine motor control required for effective toothbrushing and flossing. The formulation’s ability to reduce subgingival pathogens with no mechanical intervention suggests it may partially compensate for inadequate plaque removal, offering a valuable adjunctive therapy for individuals who struggle with conventional oral hygiene practices.

### Oral Health Disparities in the Aging Population

The study population reflects important demographic characteristics relevant to oral health disparities. Participants were predominantly female (90% overall) and racially diverse, with approximately 45% identifying as Black or African American. This composition is significant because oral health disparities persist across socioeconomic, racial, and ethnic lines in the United States, with minority populations and individuals of lower socioeconomic status experiencing disproportionately high rates of periodontal disease, tooth loss, and barriers to dental care access [49]. African American and Hispanic older adults, in particular, have been documented to experience higher prevalence and severity of periodontitis compared to their White counterparts, even after adjusting for socioeconomic factors [49]. The successful demonstration of PTx800’s efficacy in this population is particularly meaningful, as it suggests the intervention may help address oral health disparities by providing an accessible, effective option for communities that face barriers to traditional dental care. The lozenge format requires no prescription, specialized equipment, or professional administration, making it a feasible option for integration into community health programs, senior centers, or public health initiatives targeting underserved older adults.

### Baseline Health Characteristics and Study Robustness

A notable feature of this trial was the imbalance in self-reported baseline systemic health conditions between groups. The PTx800 group had a substantially higher burden of conditions including hypertension (75% vs. 30% in controls), arthritis (43% vs. 13%), and dry mouth complaints (32% vs. 0%). These conditions are known to negatively impact oral health through multiple pathways. Arthritis impairs the manual dexterity required for effective toothbrushing and flossing, directly compromising mechanical plaque removal [48].

Many antihypertensive medications cause xerostomia, reducing the protective effects of saliva and increasing susceptibility to plaque accumulation and gingival inflammation. Dry mouth itself is associated with increased risk of dental caries, candidiasis, and periodontal disease due to diminished antimicrobial proteins and buffering capacity [41,43].

Despite participants in this group having a greater baseline health disadvantage, PTx800 demonstrated significant clinical and microbiological improvements. This strengthens confidence in the intervention’s efficacy and suggests that PTx800 can be effective even in populations with comorbidities that typically predict poorer oral health outcomes. This finding has important clinical implications, as older adults with multiple chronic conditions are a population in need of accessible oral health interventions that can be effective despite physical limitations.

### Study Limitations

Several limitations must be acknowledged when interpreting these findings. First, participant recruitment and retention proved challenging. This may have been due to our strict inclusion criteria alongside the documented challenges in recruiting recruitment and retention of older adults in clinical trials [50]. Consequently, the small sample size (n=20 total; 11 intervention, 9 control) reduces statistical power and limits the generalizability of the results. Nevertheless, because older adults are underrepresented in clinical trials [51], generating preliminary safety and efficacy data in this population provides valuable groundwork for larger, fully powered follow up studies. While the statistically significant differences between groups are encouraging, the study would benefit from replication in larger cohorts to confirm these findings and enable subgroup analyses by demographic characteristics, baseline disease severity, or comorbidity profiles. The six-week study duration, while sufficient to detect meaningful changes in inflammatory markers and microbial composition, may not be able to detect the long-term effects on attachment loss, bone resorption, or tooth retention. Extended follow-up studies are needed to determine whether the observed improvements are sustained over time and whether they translate into clinically meaningful reductions in periodontitis progression or tooth loss.

The imbalance in baseline systemic health conditions, while potentially strengthening the case for PTx800’s efficacy as discussed above, introduces confounding variables that complicate interpretation. Future trials should stratify randomization by key health conditions or employ propensity score matching to ensure more balanced groups. The study was conducted in Northeast Ohio with senior center participants, which limits generalizability to other geographic regions, cultural contexts, or populations with different oral health literacy levels or access to dental care.

While this study assessed clinical and microbiological outcomes, it did not measure potential systemic effects such as changes in circulating inflammatory markers, cognitive function, or cardiovascular parameters. Given the proposed associations between periodontal pathogens and systemic diseases, future studies incorporating these measures would provide valuable insights into whether oral health improvements translate into systemic health benefits.

### Clinical Implications

The results of this trial have practical implications for clinical practice and patient care. PTx800 lozenges represent a promising adjunctive therapy for older adults with gingivitis, particularly those with physical limitations, xerostomia, or other conditions that impair conventional oral hygiene practices. The lozenge format offers ease of use and direct delivery of bioactive ingredients throughout the oral cavity including difficult-to-reach subgingival sites.

For dental practitioners, PTx800 could be recommended as part of a comprehensive oral health maintenance strategy, particularly for patients who struggle with plaque control despite instruction in proper brushing and flossing techniques. The intervention may be especially valuable during the early stages of gingivitis when intervention can prevent progression to periodontitis and irreversible tissue damage. Given the active placebo control design, clinicians can be confident that PTx800 provides benefits beyond sugar alcohols alone.

For public health practitioners and policymakers, PTx800’s demonstrated efficacy in an underserved population suggests potential applications in community-based oral health programs targeting health disparities. The intervention’s accessibility and ease of use make it suitable for distribution through community health centers, senior centers, or other public health initiatives, potentially reaching older adults who face barriers to traditional dental care.

The observed reduction in periodontal pathogens, particularly *P. gingivalis*, raises the possibility that interventions targeting oral health may have broader health implications for older adults. While this study did not assess systemic health outcomes, the documented associations between periodontal pathogens and conditions such as Alzheimer’s disease and cardiovascular disease suggest that managing oral pathogen burden could be an overlooked strategy for systemic disease prevention. This merits further study in geriatric medicine and preventive health care, potentially elevating oral health from a quality-of-life concern to an integral component of chronic disease management.

### Implications for Future Research

The findings of this study open several important avenues for future investigation. First, larger, multicenter randomized controlled trials with extended follow-up periods are needed to confirm PTx800’s efficacy, assess the durability of microbiome modulatory effects, and evaluate whether clinical improvements are maintained with continued use or after discontinuation. Such studies should include diverse populations across age ranges, geographic regions, and health status profiles to establish generalizability.

Second, mechanistic studies are warranted to elucidate the specific pathways through which PTx800 exerts its antimicrobial and anti-inflammatory effects. In vitro studies examining PTx800’s effects on periodontal pathogen growth, biofilm formation, virulence factor expression, and host inflammatory responses would provide valuable insights. Animal models of periodontitis could assess tissue-level effects and mechanisms of action in vivo.

Third, given the strong associations between *P. gingivalis* and systemic diseases, particularly Alzheimer’s disease and cardiovascular disease, future trials should investigate whether sustained reduction of this pathogen through PTx800 supplementation influences systemic disease outcomes. Studies in populations at risk for cognitive decline could assess whether PTx800 reduces circulating inflammatory markers, slows progression of cognitive impairment, or decreases biomarkers of neuroinflammation. Similarly, trials in individuals with cardiovascular risk factors could evaluate whether oral pathogen reduction translates into improved endothelial function, reduced atherosclerotic progression, or favorable changes in cardiovascular inflammatory markers.

Fourth, implementation studies are needed to understand how PTx800 could be integrated into public health programs, community health centers, or dental practices serving older adults, particularly in underserved communities. Cost-effectiveness analyses comparing PTx800 supplementation to standard periodontal therapies would inform policy decisions and insurance coverage considerations.

Finally, studies examining the oral microbiome composition at a deeper taxonomic resolution using metagenomic sequencing would provide insights into whether PTx800 selectively reduces pathogenic species while preserving or promoting beneficial commensal bacteria, which would support its characterization as a microbiome-modulating rather than a broadly antimicrobial intervention.

## CONCLUSIONS

In conclusion, this randomized controlled pilot trial provides compelling preliminary evidence that PTx800 lozenges significantly improve clinical and microbiological markers of gingival health in older adults with gingivitis. The intervention achieved complete elimination of BOP, stabilized pocket depth, and substantially reduced red complex periodontal pathogens, outperforming an active sugar alcohol placebo control. These findings are noteworthy given the study population’s elevated systemic disease burden and the critical role of oral health in the aging population. The reduction of *P. gingivalis* and other red complex bacteria may have implications extending beyond oral health, given these pathogens’ associations with Alzheimer’s disease, cardiovascular disease, and other systemic conditions. PTx800 represents a promising, accessible intervention for managing gingivitis in older adults, particularly those in underserved communities or with physical limitations that impair conventional oral hygiene. Further research is needed to confirm these findings in larger populations, investigate long-term outcomes, and explore potential systemic health benefits of reducing periodontal pathogen burden.

## ACKNOWLEDGMENTS

We would like to thank all of the staff at Benjamin Rose for their assistance in the execution of this clinical study and Ashley Beiniek for helping to organize the blinded data sets.

## DATA AVAILABILITY

The data that support the findings of this study are available from the corresponding author upon reasonable request.

